# Building a community-academic partnership in public housing: Lessons from applying the CBPR Partnership Readiness Model

**DOI:** 10.64898/2026.10.28.26366741

**Authors:** Nadia A. Sesay, Kendra OoNorasak, Jayden E. Brewer, Mansura Shahad Bawa, Dana Mason, Kristin Hughes, Jessica Coffie, Brandi M. White

## Abstract

The DASH (Dietary Approaches to Stop Hypertension) eating plan has been shown to reduce hypertension; however, adherence is low among low-income African American (AA) women, who experience a disproportionate burden of hypertension. To better understand barriers to DASH among low-income AA women, specifically AA women living in public housing, we established a community-academic partnership using a project advisory board (PAB). This paper applies the CBPR Partnership Readiness Model to assess community stakeholder readiness for the partnership. PAB members were recruited through ongoing research and volunteer work in the target community. We developed a 33-page PAB workbook clarifying study goals, member roles and expectations, and hypertension information. A questionnaire was used to gather feedback on members’ experiences. PAB membership increased by 150% after the first year following a second recruitment phase. Members reported satisfaction with progress toward local food equity initiatives and with partnership sustainability. Findings indicate the model was helpful in evaluating the PAB formation process for a project involving public housing stakeholders. PABs can help build local support for community-engaged projects. When members can see the advantages of their involvement, partnerships are strengthened.

## Introduction

One strength of community-academic partnerships is the coupling of scientific and locally produced knowledge to raise awareness and understanding of chronic disease risk factors in medically underserved communities (Wallerstein & Duran, 2006). These partnerships, when developed within a community-based participatory research (CBPR) paradigm, are also shown to increase acceptance of evidenced-based interventions (Chassler et al., 2025). A project advisory board (PAB) further supports the partnership by enhancing community representation, garnering local support for the project, and steering project functions (Ortega et al., 2018). Best practices for community-based participatory research (CBPR) suggest that a PAB is assembled with intent; potential members are carefully identified by criteria like affinity or residence, ongoing prosocial work within and for the target community, and shared illness experience (Newman et al., 2011).

Assessing readiness between community stakeholders and the academic team is a critical step in developing the PAB. Readiness is determined by mutual understanding of community centered values, study aims and research operations as misalignment can endanger partnership sustainability and hinder achievement of project goals (Andrews et al., 2012). Studies explain promising approaches for ensuring readiness within partnerships that result in the reduction of health disparities in minority groups (Chassler et al., 2025; Newman et al., 2011; Wallerstein & Duran, 2006; Whitt-Glover et al., 2013). Thus, we applied the CBPR Partnership Readiness Model (Andrews et al., 2012) to determine community stakeholder readiness to form the PAB for a multiyear community-based study developing a dietary hypertension intervention for African American (AA) women living in subsidized housing.

### Public Health Concern

AA women have significantly higher estimates of hypertension compared to women of all other race and ethnic groups, resulting in a disproportionate burden of stroke and heart disease (Martin et al., 2025). Racial disparities in cardiovascular disease are largely attributed to poor social determinants of health, including chronic stress, barriers to accessing quality healthcare, and increased socioeconomic barriers (Carnethon et al., 2017; Ferdinand et al., 2020). Economically-disadvantaged AA women, especially those living in public housing or subsidized housing, are at an even higher risk for hypertension because they are less likely to be aware of hypertension and other cardiovascular disease risk factors (Al-Bayan et al., 2016; Behara et al., 2000; Ford et al., 2009; Spikes et al., 2022).

Diet is a crucial modifiable risk factor for preventing and managing hypertension and reducing health disparities (Arnett et al., 2019). Adopting the DASH (Dietary Approaches to Stop Hypertension) eating plan is one evidence-based strategy to significantly reduce blood pressure and hypertension risk, and improve overall cardiovascular health (Filippou et al., 2020; Guo et al., 2021). The DASH plan emphasizes eating heart-healthy foods (e.g., fresh vegetables, fruits, whole grains, and lean meats) and limiting sodium intake, saturated fats, and sugary drinks and sweets (National Heart, Lung, and Blood Institute, 2025).

While there is evidence to demonstrate AAs and women who adhere to the DASH eating plan experience greater cardiovascular benefits compared to their White and male counterparts (Jeong et al., 2023), DASH adoption/adherence among AAs remains low, especially among low-income populations (Bertoni et al., 2011; Epstein et al., 2012; Whitt-Glover et al., 2013). Barriers to DASH adoption for low-income AA communities include limited access to culturally relevant DASH recipes (e.g., soul food) and affordable DASH-friendly foods, as well as a lack of adequate kitchen supplies for DASH meal preparation (White et al., 2024). Given the high rates of hypertension among AA women and the established benefits of DASH, there is a need for culturally responsive strategies to increase DASH adoption in this population, especially for low-income AA women such as those living in public housing. In response, academic partners developed *DASH to the Table*, a project that aims to create a low-cost, soul food DASH cookbook in collaboration with AA women in public housing and key stakeholders to promote DASH adoption.

### Community Engagement and Formative Work

This project builds on long-standing, community-engaged efforts to address social determinants of health in Lexington, Kentucky’s East End, a historically AA community. Since 2018, members of the academic team have partnered with the Lexington-Fayette Urban County Housing Authority (LHA), which manages the city’s public housing, to build trust, identify community residents’ health priorities, and support health promotion initiatives. With support from an intramural grant, the academic team conducted formative research to understand barriers to cardiovascular health and identified limited access to healthy, affordable food as major barriers (Smith & White, 2021; Sullivan et al., 2025). In response, the Health Equity Action Team (HEAT) was established in 2019 to address community concerns and guide a community-driven research agenda for academic partners. HEAT included AA women who were public housing residents, LHA staff, college students, public health officials, and other key stakeholders. Meetings fostered trust and fellowship and were often facilitated by sharing heart-healthy meals, which sparked interest among residents in adapting traditional recipes to better align with heart-healthy eating plans such as DASH. These discussions informed the development of the *DASH to the Table* grant application.

In addition to relationship-building with the community through HEAT, members of the academic team had previous collaborative research and volunteering experience with stakeholders committed to food security efforts and nutrition education. One member of the academic team. co-founded and spearheaded a campus-based organization that strives to address food waste and insecurity across the Lexington community, including families with children, older adults, and college students, through sustainable approaches, educational resources, and mutually beneficial partnerships with stakeholders who later became members of the PAB. The relationship between the academic partner and a particular member of the PAB has been nurtured for five years through various grant and research projects prior to the *DASH to the Table* project. Another member of the academic team volunteered with local organizations distributing free, healthy cooked meals and providing farmers’ market produce at affordable prices for low-income community members and later joined the project as a research assistant. A resident of one community identified for collaboration in *DASH to the Table* also joined the project as a research assistant.

### CBPR Partnership Readiness Model

Existing relationships between the academic team and key stakeholders facilitated the identification and recruitment of potential PAB members. We applied domains from the CBPR Partnership Readiness Model (Figure 1) to determine mutual readiness to commence the partnership (Andrews et al., 2012). Readiness entails alignment of mutual values, understandings of responsibilities, and expectations from both partners. Readiness is also iterative, as assessment of the community-academic partnership across dimensions of the partnership readiness model is ongoing (Andrews et al., 2011; Andrews et al., 2012). We describe the recruitment, formation, and early implementation of the PAB and evaluate this process using the model.

**Figure 1.**
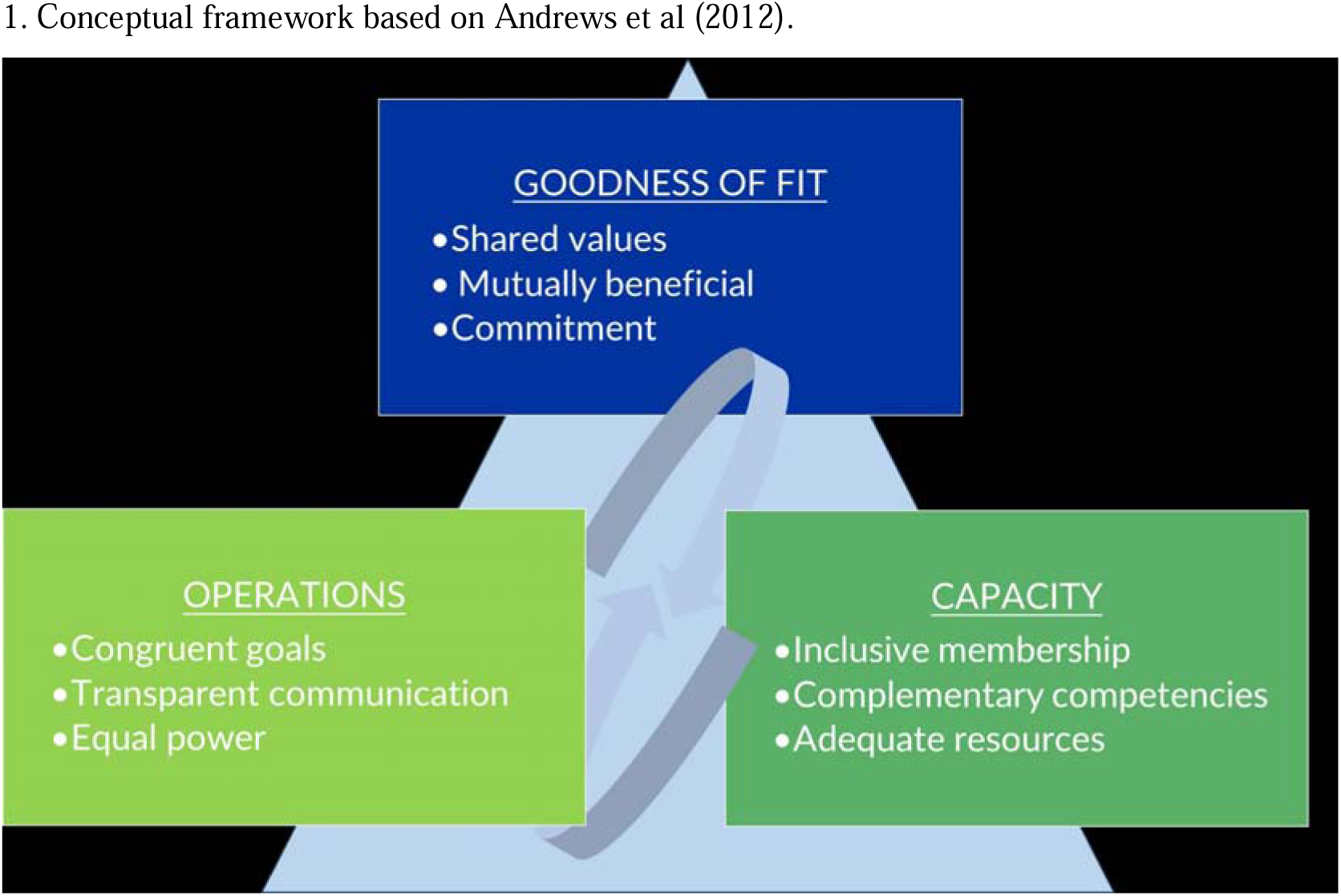
CBPR Partnership Readiness Model Domains Applied to PAB Formation^1^.

## Materials and Methods

### Study Context and Design

*DASH to the Table* is guided by the Gelberg-Anderson Behavioral Model for Vulnerable Populations, which highlights predisposing factors (e.g., limited economic resources) and barriers (e.g., high-cost of DASH-friendly foods) that influence and hinder adherence to the DASH plan and contribute to higher hypertension rates among AA women living in public housing (Stein et al., 2007). The purpose of the *DASH to the Table* PAB is to discuss the best strategies to increase access to affordable DASH-friendly foods for the East End community and other local low-income communities at risk for hypertension in Lexington. The PAB is composed of community stakeholders who share an interest in the aims of the research project and include AA women living in public housing, housing authority staff, community-based organizations, and food and nutrition security advocates. Members provide insight and personal experiences to study implementation (Newman et al., 2011; Ortega et al., 2018). Their participation broadens the scope of perspectives, while also raising awareness of often-overlooked health issues and disparities (Newman et al., 2011; Ortega et al., 2018). PABs foster trust with academic partners, which encourages research participation among historically marginalized groups (Ortega et al., 2018), a component especially crucial when working with AA communities.

Using a multi-phase mixed methods design, the project includes key informant interviews with the target population to understand DASH adoption barriers, recipe adaptations and evaluations, and community-based DASH cooking sessions. The PAB was established to guide each phase of the study to ensure the project is responsive to community needs and preferences. This paper reflects our work from January 2024 to June 2025. All study procedures were approved by the University of Kentucky’s Institutional Review Board (IRB #90193).

### Community of Interest

LHA manages public housing and provides affordable housing opportunities for low- and moderate-income individuals and families (LHA, n.d.). Of Lexington’s 2,168 public housing residents, 81% are AA, 94% live in poverty, and 86% have a female head of household (U.S. Department of Housing and Urban Development, 2026). The study’s inclusion criteria are at least 18 years of age, identify as a Black/AA woman, and residing in public housing/receiving Section 8 housing vouchers or any other form of housing assistance in Lexington. Thus, with the support of LHA, we recruited participants from a community in Lexington’s East End managed by the organization.

### PAB Recruitment and Membership

The timeline for recruiting and establishing the PAB is shown in Table 1. After the grant was awarded, the academic team met with potential key stakeholders for the PAB between January and April 2024 to determine mutual interest in the project aims. We operationalized key stakeholders as community members from the target population (i.e., AA women living in public housing), individuals associated with the local housing authority, and/or local food/nutrition security advocates. Key stakeholders that indicated an interest in joining the PAB were sent the eligibility survey and informed consent document using REDCap (Research Electronic Data Capture), a secure, web-based platform designed to support data collection and management for research studies (Harris et al., 2009). Eligibility for the PAB was individuals 18 years old or older with an interest in increasing access to healthy foods, improving the health of AA women, or improving the health of public housing residents in the local community. Those who met eligibility criteria and completed informed consent were invited to formally join the PAB.

**Table 1.** Timeline of PAB Recruitment and Formation.

| Date | Action |
|---|---|
| November 2018 – *Ongoing* | Community engagement and formative work |
| February 2023 | Grant submitted with letter of support from housing authority |
| December 2023 | Grant awarded |
| **PAB Phase 1** |  |
| January – April 2024 | Met with potential PAB members to gauge interest (Recruitment) |
| May – June 2024 | PAB invitations emailed<br>Electronic informed consent completed<br>Distributed PAB Workbook to members |
| July 2024 | PAB meeting 1 (virtual) |
| August 2024 | PAB meeting 2 (virtual) |
| September 2024 | PAB meeting 3 (virtual) |
| October 2024 | PAB meeting 4 (virtual) |
| November – December 2024 | Additional recruitment initiated based on PAB feedback |
| **PAB Phase 2** |  |
| December 2024 | PAB invitations emailed (Recruitment of additional members)<br>Electronic informed consent completed<br>Distributed PAB Workbook to members |
| January 2025 | PAB meeting 5 (in person) |
| June 2025 | PAB meeting 6 (virtual) |

The academic team initially identified ten key stakeholders and invited them to meet to discuss the project and PAB interest. Eight stakeholders agreed to meet and all expressed interest in joining the PAB. We emailed these stakeholders the eligibility survey and informed consent form and received a recruitment response rate of 75% (6 of 8 invited stakeholders) (percentage of persons who consented after being invited). Among those who consented, the PAB participation rate was 67% (4 of 6 stakeholders consented and attended at least one PAB meeting).

Based on feedback from early PAB meetings (July - October 2024), members recommended expanding the board to increase representation from the project’s target population. In response, the academic team resumed PAB recruitment in November - December 2024. This second recruitment phase focused on engaging new community members and local food/nutrition security advocates not reached during initial recruitment, as well as re-engaging community members who had previously expressed interest but had not completed the consent process or had consented but not participated. During this phase, we met with six interested stakeholders, all of whom expressed interest, completed informed consent (100% recruitment response rate), and attended at least one PAB meeting (100% participation rate). This brought total PAB membership to ten individuals. Demographic characteristics and affiliations of PAB members are provided in Table 2.

**Table 2.** PAB Characteristics and Organizational Affiliations.

| Characteristic | PAB Total<br>(n=10) | PAB Phase 1<br>(n=4) | PAB Phase 2<br>(n=6) |
|---|---|---|---|
| Mean age in years (standard deviation) | 44.33 (13.5) | 44.75 (8.7) | 44.00 (17.5) |
| Female, % | 90% | 100% | 83% |
| African American/Black race, % | 90% | 75% | 100% |
| Affiliation*, %: |  |  |  |
| Community member | 30% | 0% | 50% |
| Community-based organization | 50% | 75% | 33% |
| Housing authority | 30% | 25% | 33% |

*\*Note:* Categories are not mutually exclusive; some individuals held multiple roles.

### PAB Workbook

We created a 33-page PAB workbook that expounded on the triad dimensions of the CBPR Partnership Readiness Model. Specifically, the workbook in plain language described the study’s goals, expectations, and key concepts related to hypertension and the DASH plan. The workbook served as an orientation to the project and a capacity-building resource. We developed the workbook using Canva, an online graphic design platform that facilitates creation of mixed media presentations. Using the image archive in Canva, we incorporated soul food iconography into the workbook (e.g., okra and black-eyed peas) to visually convey the project aims and underscore cohesion in our messaging with the PAB. We distributed the workbook electronically to the PAB prior to the first meeting so that members could familiarize themselves with its contents. Each member also received a printed workbook to support their understanding and ongoing participation. New members recruited during the second recruitment phase also received the workbook. For new members recruited during the second recruitment phase, their first meeting also served as an orientation and refresher for existing members.

### Meeting Procedures and PAB Member Assessment

The academic team coordinates and facilitates all PAB meetings, and PAB members receive a $50 gift card for each meeting attended. Scheduling the meeting begins approximately 2 to 4 weeks before the estimated meeting date (Figure 2). Email is the primary and PAB-preferred mode of communication. A total of four two-hour meetings have been/will be held per year over the three-year project period and are held either in person or via Zoom, based on group preference. Heart-healthy meals are served at in-person meetings. To document and assess PAB formation and implementation, the academic team records and transcribes all meetings and tracks meeting attendance (Table 3). All transcripts are de-identified. An agenda is distributed before each meeting, and meeting minutes are shared with PAB members within two weeks following the meeting. Feedback on the minutes from PAB members is requested prior to the finalizing of minutes.

**Figure 2.**
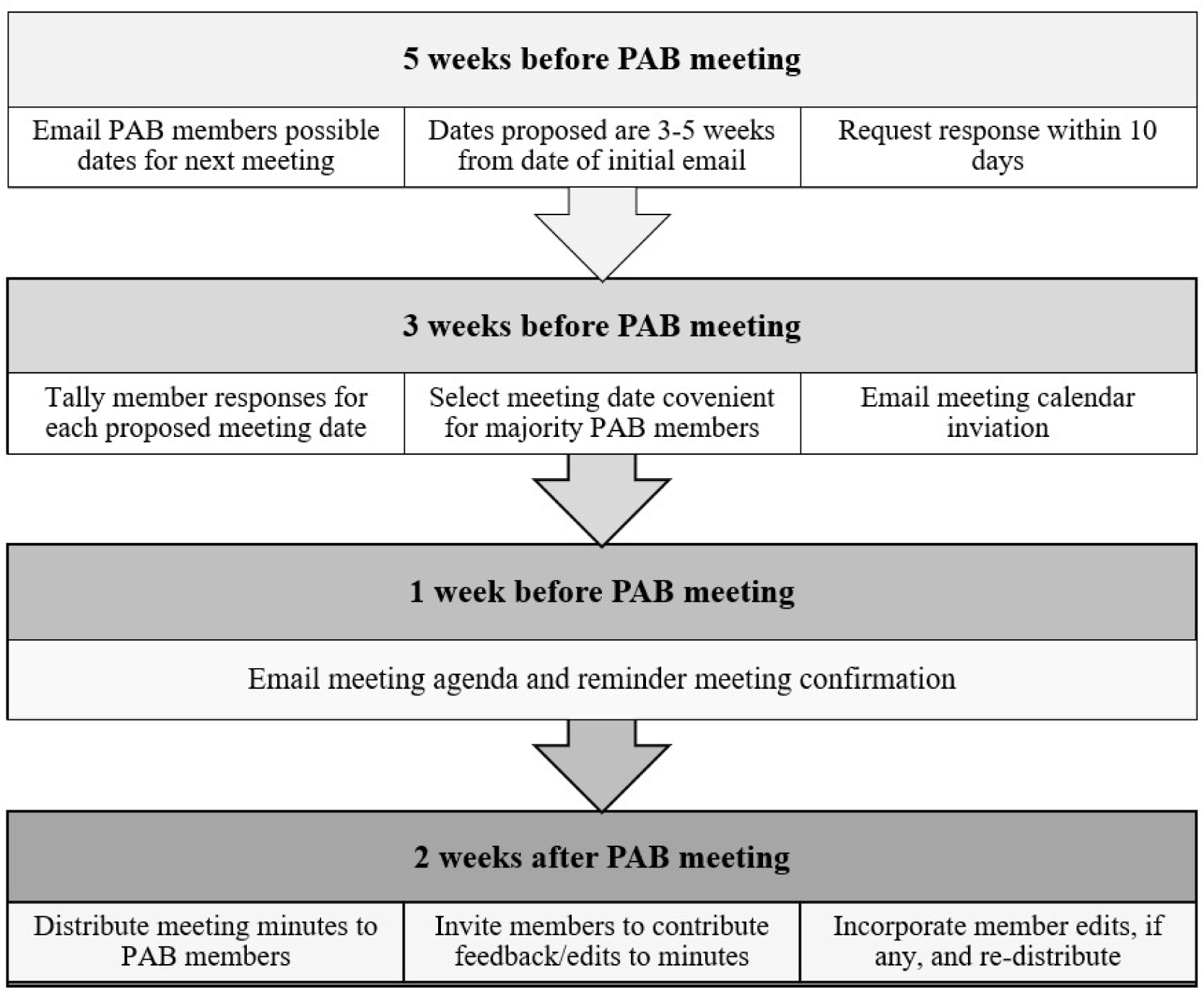
Activities and Timeline for Coordinating PAB Meetings.

**Table 3.** PAB Meeting Attendance by Phase.

| **PAB Meeting** | **Members Present** |
|---|---|
| PAB Phase 1, (n=4): |  |
| PAB 1 | 75% |
| PAB 2 | 100% |
| PAB 3 | 50% |
| PAB 4 | 75% |
| Mean attendance rate | 75% |
| PAB Phase 2, (n=10): |  |
| PAB 5 | 90% |
| PAB 6 | 40% |
| Mean attendance rate | 65% |

### Partnership Readiness: Data Sources and Assessment

We also seek to understand PAB members’ experiences working in partnership with the academic team. We administer a brief REDCap survey to PAB members at the end of each project year. The year-end survey administered to Phase 1 members in December 2024 utilized open-ended questions to record members’ experience across multiple categories of PAB functioning (Table 4). The survey is anonymous, allowing for candid feedback on potential areas of improvement that could enhance the community-academic research relationship. In addition, the academic team holds post-PAB meeting debriefs to reflect on member engagement, communication dynamics, and barriers to participation.

**Table 4.**
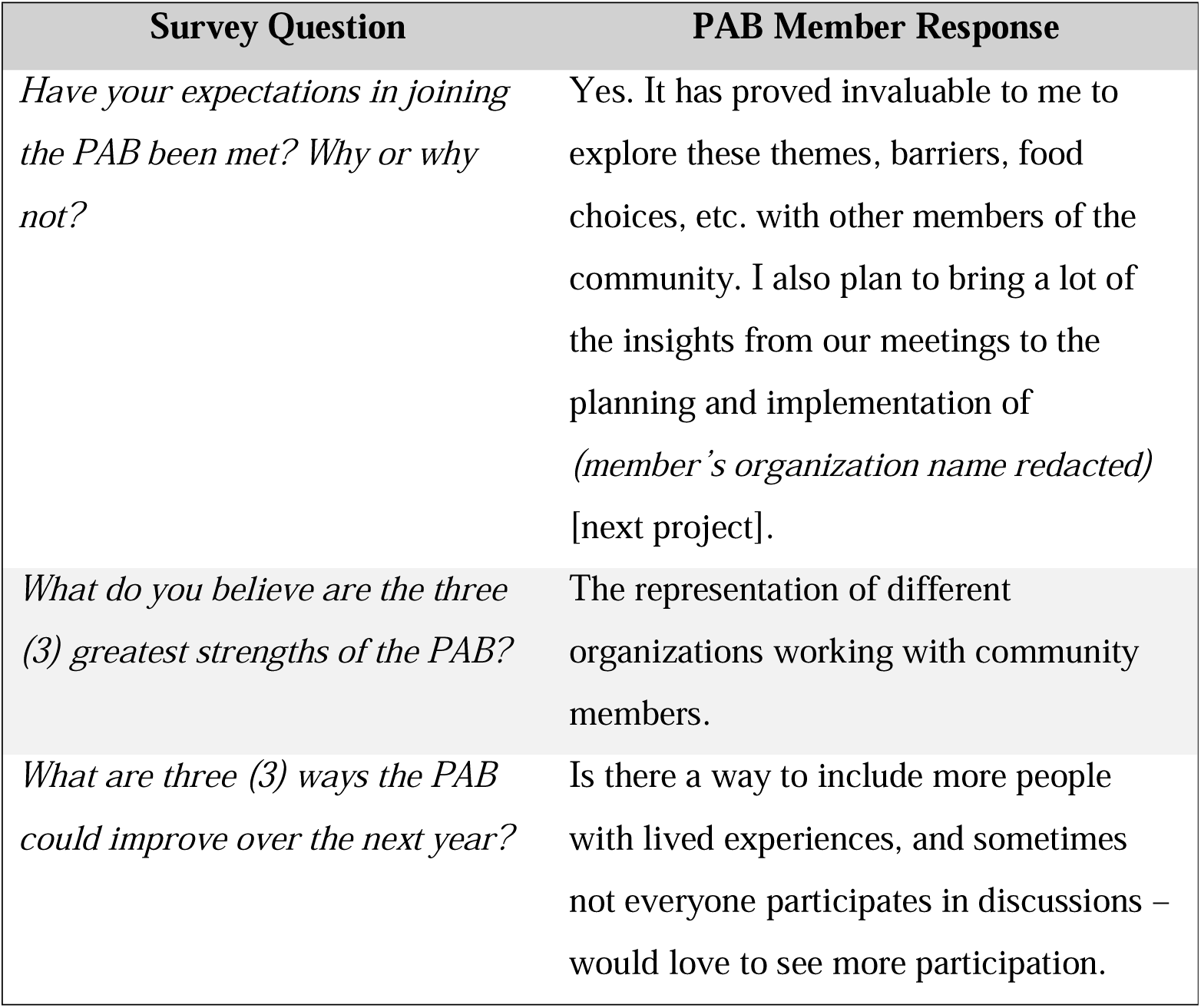
PAB Member Feedback on Partnership Experience.

We used the CBPR Readiness Model (Figure 1) to evaluate the PAB process by examining three domains: Goodness of Fit, Capacity, and Operations. Goodness of Fit explored alignment between PAB member interests and the project’s goals. Capacity focused on PAB member confidence contributing to meeting discussions and additional project-related needs. Operations examined meeting logistics, communication preferences, and meeting attendance. While a formal analysis was not conducted, this process helps identify partnership needs and areas for improvement to strengthen the PAB’s readiness and support future project implementation.

## Results and Lessons Learned

From Phase 1 to Phase 2, PAB membership increased 150%. We maintained 75% PAB member attendance and 65% member attendance at PAB meetings in Phase 1 and Phase 2, respectively (Table 3). Open-ended survey responses showed that meeting discussions provided inspiration in steering their respective organizations towards food equity and sustainability projects and an appreciation for the diversity of the membership group. Members also provided suggestions on diversifying future PAB participation (Table 4).

An evaluation of the recruitment, formation, and early implementation of the PAB using the CBPR Partnership Readiness Model provides insights into building and sustaining community-academic partnerships with public housing communities. Reflecting on our experiences allowed us to identify strategies that supported PAB engagement and identify areas for improvement. The lessons learned summarized here may inform future efforts to establish advisory boards in similar contexts.

### Goodness of Fit

#### Shared Values

Clearly defined inclusion criteria and an initial orientation can facilitate alignment between members’ interests and the project’s mission. The PAB included individuals interested in increasing access to healthy foods, improving AA women’s health, or supporting public housing residents’ health. Community partners were engaged in collaborative efforts and had an interest or active role in health equity. The PAB workbook was an effective tool for building a shared understanding by outlining project goals, defining key concepts, and providing background on hypertension and the DASH plan. Annual evaluation survey feedback indicated the workbook was accessible and communicated the PAB’s purpose.

#### Mutual Benefit

Partnerships are strengthened when members can identify benefits to their participation. For example, one community partner is using PAB discussions to inform decisions about the types of food and meals to offer at a new grocery store in the target community. The housing authority viewed the PAB as a way to explore additional health and wellbeing resources for residents. Annual evaluation survey results showed respondents agreed their participation was valuable both to themselves and to the communities and organizations they represent.

#### Commitment

Flexible meeting formats can help address competing priorities but must also account for barriers such as limited technology access. Holding most meetings virtually allowed many members to participate despite busy schedules; however, some members with limited internet access could not attend regularly. Providing hybrid or alternative participation options could increase accessibility and sustained involvement.

### Capacity

#### Inclusive Membership

Adequate representation from the target community is important to ensuring that partnership activities reflect community priorities. Underrepresentation of community members during the first recruitment phase limited the diversity of perspectives in early discussions. A second recruitment phase was required to ensure that voices from the community of interest were represented, highlighting the importance of assessing membership composition early and adjusting when needed.

#### Complementary Competencies

Combining lived experience with professional expertise enriched PAB discussions and decision-making. Members provided cultural and contextual knowledge as AA women and public housing residents, or leaders of organizations that serve that demographic group. The academic team’s competencies covered areas like recruitment, consent, and other research operations. These complementary competencies strengthened the partnership’s capacity to discuss culturally responsive strategies to increase community access to affordable DASH-friendly foods.

#### Adequate Resources

Resources are essential to sustaining PAB engagement. Grant funding allowed for compensation of key personnel on the academic team, including research assistants, and gift card incentives for PAB members. Additional capacity was built through the involvement of undergraduate students completing independent studies in health equity and community engagement. Grant resources also supported necessary equipment for project activities. Maintaining adequate funding streams for personnel, member incentives, and equipment will be key to sustaining the PAB’s work over time.

### Operations

#### Congruent Goals

The PAB workbook outlined the academic team’s specific aims in forming the PAB, meeting operations, and conveyed how the PAB fits within overarching *DASH to the Table* goals. Members are also encouraged to share how the community-academic partnership may support their respective goals, such as in co-writing small-scale funding grants for local organizations to advance health and food equity initiatives. Furthermore, in every PAB meeting, members are invited to share information about upcoming community programs and service opportunities that the academic team and other fellow members may support.

#### Transparent Communication

The academic team confirmed the preferred communication channel with members during the first PAB meeting. As such, most communication occurs via email with occasional phone calls or text messaging for spur-of-the moment communication (i.e., that a member is running late for a meeting). Furthermore, meeting agendas are emailed ahead of the meeting and minutes are distributed following the meeting for the PAB members’ review.

#### Equal Power

The community-academic team co-designed a project logo to signal that research products disseminated to the community incorporate the input of community stakeholders. PAB members are invited as co-authors on project manuscripts and conference submissions. Open discussions about the academic institution’s power and chronic illness research within predominantly AA communities in Lexington are welcomed.

## Conclusion

We examined how the CBPR Partnership Readiness Model can be used to guide the early development of a community-academic partnership supporting DASH adoption among low-income AA women. The model was useful for evaluating the PAB formation process for a project that engaged stakeholders from the community, local housing authority, and community-based organizations. Findings from applying the model’s dimensions (Goodness of Fit, Capacity, and Operations) highlighted areas of strength and opportunities for growth, indicating the importance of reassessing the community-academic partnership as it evolves. In addition, prior engagement and trust-building were essential to partnership readiness in this public housing context. These findings provide insights for community and academic partners seeking to assess and strengthen readiness in early partnership efforts.

## Implications for Community Practice

Our findings suggest that assessing partnership readiness can be valuable for community-academic partnerships working with public housing communities. Providing accessible resources to community partners (e.g., PAB workbook) may help establish a shared understanding of project goals, roles, and expectations, which is essential in the early stages of partnership development. In addition, regularly evaluating partnership membership (i.e., Who is not represented at the table?) is important to ensure the community most impacted is included. Flexible meeting formats and regular communication were also important to supporting partnership engagement. These strategies may be helpful when establishing community-academic partnerships aimed at promoting health equity.

## Data Availability Statement

The data supporting the findings of this study are not publicly available due to privacy and confidentiality considerations associated with the small sample of community stakeholders.

## Acknowledgement

The authors express their deepest gratitude to Sandra Ballew-Barnes for her contributions to the project advisory board.

